# Lack of melatonin secretion induced by moderate hypothermia during aortic-arch surgery, possible implications in neurological outcomes

**DOI:** 10.1101/2025.11.24.25340883

**Authors:** Leone Alessandro, Occhinegro Alessandra, Piscitiello Emiliana, Paterini Paola, Bugani Simone, Mariani Carlo, Nocera Chiara, Hitrec Timna, Amici Roberto, Pacini Davide, Luppi Marco

**Author notes:** **Corresponding author:** Dr. Marco Luppi, Department of Biomedical and Neuromotor Sciences, DIBINEM, University of Bologna, division of physiology. Piazza di Porta San Donato, 2 - 40126 Bologna, Italy.

## Abstract

Preclinical data from the peculiar animal model of “synthetic torpor” (ST), a reversible hypothermic condition resembling natural torpor but pharmacologically induced in rats (a non-hibernating mammal), suggest that in hypothermic conditions the neuroprotective effects of melatonin are strongly enhanced. As a protection technique, during aortic-arch surgery patients are induced a hypothermia similar to ST. Since in ST systemic melatonin was found particularly high, our aim was to assay serum melatonin in patients (N=8) undergoing aortic surgery, either during hypothermia and in the following four recovery days. Serum markers of blood-brain barrier (BBB) integrity (the astrocytic protein S100B) and neuronal damage (neuronal-specific enolase, NSE) were also measured. Results show that, in contrast to what observed in ST, in hypothermic patients melatonin was dramatically reduced in respect to the pre-anesthesia level, slowly recovering during the post-surgery period. Also, S100B and NSE raised during surgery, indicating highly leaking BBB and some ongoing neuronal damage, though both markers returned closer to pre-anesthesia levels within recovery period. Together, present results show that, in aortic-arch surgery, hypothermic patients totally lack the systemic melatonin peak that was observed in ST, the BBB was temporarily compromised and some acute neuronal damage occurred. A main implication of this work is that, exploiting the low BBB efficiency that makes easier to reach brain parenchyma, by administering melatonin during the hypothermic stage of the surgery the observed gap could be filled, possibly triggering the neuroprotective mechanism seen in preclinical observations and leading to better neurological outcomes for this surgical procedure.

**Highlights:**

- During hypothermia induced in aortic-surgery systemic melatonin was low
- Serum biomarkers enlighten transient blood-brain barrier and neuronal sufferings
- Giving melatonin during aortic-surgery hypothermia may improve neurological outcome

## Introduction

It is well known that melatonin has neuroprotective and antioxidant effects on the nervous system, strongly enough that it has been seriously considered for developing possible treatments to contrast neurodegenerative diseases and dementias (Herrera-Arozamena et al., 2016; Shukla et al., 2017). Unfortunately, different clinical trials conducted in recent years did not confirm the usefulness of melatonin in coping with cognitive impairments (Sanchez-Barcelo et al., 2017). However, recent preclinical data on rodents, obtained from our laboratory (Squarcio et al., 2023), opened the possibility to reconsider the neuroprotective function of melatonin for clinical use. Specifically, by using the “synthetic torpor” (ST) animal model (e.g., a transient and reversible hypothermic condition resembling hibernation, but pharmacologically induced on rats) (Cerri et al., 2013; Cerri, 2017), it was found that, surprisingly, melatonin is abundantly secreted during the hypothermic phase and this bulk secretion seems to act on neurons by triggering a protective molecular mechanism (Squarcio et al., 2013; Hitrec et al., 2024). Even though the effects of this mechanism were only partially investigated, apparently it seems to contrast neuronal suffering by deactivating glycogen-synthase kinase 3-β (GSK3β) and the accumulation and aggregation of hyperphosphorylated Tau protein (Luppi et al., 2019; Hitrec et al., 2021; Squarcio et al., 2023; Hitrec et al., 2024), one of the major specific hallmarks observed in a broad class of neurodegenerative diseases, defined as “tauopathies” (Wang and Mandelkow, 2016), and also involved in neuronal damage following ischemia (Pluta et al., 2021). It is worth noting that this protective mechanism was triggered by the ST and starts right during the hypothermic condition, not only when animals recovered euthermia (Squarcio et al., 2023). The suggested interpretation by the authors was that melatonin, when acting on neurons at low body temperature, may express some peculiar functioning in a way that is not possible at physiological temperature (Squarcio et al., 2023; Hitrec et al., 2024). This may also explain the reason why clinical trials using melatonin as a neuroprotective agent were not as successful as expected (Sanchez-Barcelo et al., 2017).

The not obvious link between ST and cardiac surgery is based precisely on hypothermia, induced in patients with the hypothermic circulatory arrest, a strategy used as cerebral and visceral organ protection technique during aortic arch surgery (Pacini et al., 2007; Pacini et al., 2014). When it was first introduced, hypothermic circulatory arrest was performed at temperatures of about 18 °C (Borst e al., 1983; Kazui et al., 1994). The development of further cerebral protection strategies such as antegrade selective cerebral perfusion allowed a progressive shift towards higher temperatures with good results in terms of mortality and neurological complications, and mild-to-moderate hypothermia with a nasopharyngeal temperature of 25-26 °C is now the most widespread technique (Kamiya et al., 2007; Urbanski et al., 2012). This, along with the development of surgical techniques such as the Elephant Trunk and, more recently, the introduction of branched grafts simplified the treatment of complex pathologies of the thoracic aorta.

Since, during ST, hypothermia by itself seems to represent a stimulus strong enough to trigger the melatonin bout, here we hypothesized that the same process should be observed in humans subjected to hypothermia during the aortic surgery, considering that the brain temperature is very similar in the two conditions.

Hence, to verify this hypothesis we determined melatonin systemic levels in patients undergoing aortic arch surgery with the use of moderate hypothermia at 25°C of nasopharyngeal temperature as cerebral and visceral organ protection method, during aortic arch replacement.

Moreover, to understand whether systemic melatonin may act easily on the brain parenchyma, we also determined systemic levels of S100B, as a marker of the blood-brain barrier (BBB) integrity (Marchi et al., 2003; Sandroni et al., 2023), and neuronal-specific enolase 2 (NSE), as a marker of BBB integrity and neuronal damage (Marchi et al., 2003; Sandroni et al., 2023).

## Methods

Between January 2021 and March 2022, a total of 8 patients underwent total aortic arch replacements in the department of Cardiovascular Surgery, S. Orsola Hospital, University of Bologna, Italy. The patients’ characteristics are reported in Table 1.

**Table 1:** pre- and intra-operative characteristics.

|  | N=8 | N=8 |
| --- | --- | --- |
| Mean age (n, SD) | 59 | 7.9 |
| Male sex (n, %) | 5 | 62.5% |
| BMI (n, SD) | 25.9 | 3.4 |
| Etiology |  |  |
| Degenerative aneurysm (n, %) | 5 | 62.5% |
| Post-dissection Aneurysm (n, %) | 3 | 37.5% |
| Renal failure (n, %) | 1 | 12.5% |
| Coronary disease (n, %) | 1 | 12.5% |
| Diabetes (n, %) | 2 | 25% |
| Smoking (n, %) | 3 | 37.5% |
| COPD (n, %) | 2 | 25% |
| Hypertension (n, %) | 7 | 87.5% |
| Connective tissue disease (n, %) | 1 | 12.5% |
| Previous cardiac surgery (n, %) | 4 | 50% |
| Associated procedures (n, %) | 4 | 50% |
| CABG (n, %) | 1 | 12.5% |
| AVR (n, %) | 1 | 12.5% |
| Bentall (n, %) | 2 | 25% |
| Cannulation site |  |  |
| BCT (n, %) | 4 | 50% |
| Axillary artery (n, %) | 3 | 37.5% |
| Ascending aorta (n, %) | 1 | 12.5% |
| CPB time (minutes, SD) | 223.6 | 42.4 |
| Cross-clamp time (minutes, SD) | 130 | 47.7 |
| ASCP time (minutes, SD) | 79.8 | 32.4 |
| VIT (minutes, SD) | 26.9 | 5.3 |
| Preoperative ELD positioning (n, %) | 5 | 62.5% |

SD: Standard Deviation; BMI: Body Mass Index; COPD: Chronic Obstructive Pulmonary Disease; CABG: Coronary artery bypass grafting; AVR: Aortic valve replacement; BCT: Brachio-cephalic Trunk; CPB: Cardio-pulmonary Bypass; ASCP: Antegrade selective cerebral perfusion; VIT: Visceral Ischemia Time; ELD: Epidural Liquor Drainage.

All elective patients underwent routine pre-operative exams, including coronary angiography (if aged 50 years or older), echocardiography and computer tomography scans (CT scan).

### Serum markers measurement timing

The measurement timing for serum levels of melatonin, S100B and NSE was managed by the anesthesiologist during the initial phase of the operation and the following post-operative days as listed below:

- ti: induction phase of general anesthesia
- t1: target temperature of 25°C
- t2: chest closure;
- G1: first post-operative day
- G2: second post-operative day
- G3: third post-operative day
- G4: fourth post-operative day

Blood samples withdrawals were subjected to standard procedures to separate serum. Then, samples were aliquoted and stored at -80 °C until assays.

### ELISA determinations

The determinations of the planned biomarkers were conducted on different aliquots of the same samples of serum. Concentration of melatonin was measured by Human MT (Melatonin) ELISA Kit (E-EL-H2016-96T; CliniSciences). While, concentrations of S100B and NSE were measured by Luminex, using a custom-developed panel from R&D Systems (LXSAHM-02). For all the ELISA procedures, the manufacturer instructions were precisely followed.

### Statistical analysis

Statistical analysis was carried out using SPSS software (28.0). ELISA results for melatonin were analyzed with a 2-way ANOVA with repeated measures. Then, means of the different observational conditions were compared with the “ti” condition, taken as the baseline value, using the modified *t-*test (*t*\*) (Holm, 1979) and correcting the α level using Bonferroni’s method. Since ELISA results for S100B and NSE were not normally distributed (Shapiro-Wilks test was significant for both), the statistical analysis for these data were conducted using a non-parametric test strategy: firstly, a Friedman test was conducted on the whole set of data, then, being significant, group means were compared with the baseline (i.e., ti condition) using the Wilcoxon test.

### Surgical technique

All operations were performed through a full sternotomy. After systemic heparinization, Cardiopulmonary bypass was instituted using the right axillary artery, femoral artery, innominate artery or ascending aorta as an arterial cannulation site. Right atrial cannulation was used to ensure venous drainage. A left ventricular vent was inserted through the right superior pulmonary vein. Core cooling to 25°C nasopharyngeal temperature was then accomplished, and circulatory arrest was instituted. Myocardial protection was achieved with infusion of crystalloid cardioplegia (Custodiol; Koehler Chemie, Alsbach-Haenlein, Germany). The aortic arch was opened, and bilateral antegrade selective cerebral perfusion was initiated by clamping the brachiocephalic trunk and cannulating the right carotid artery, in case of axillary or brachiocephalic trunk cannulation only. In order to guarantee a flow rate of 10 mL/kg/min, flow was uniformly distributed to the right and left cerebral hemispheres and adjusted to maintain the right radial pressure between 40 and 80 mmHg. Near Infrared Ray Spectroscopy (NIRS) was used to monitor cerebral perfusion during the procedure. After the arch was completely resected and removed, the proximal thoracic descending aorta was inspected and the presence of a large intimal tear was documented at the convexity of the aortic arch at the isthmus level. The lesion was confirmed by the angioscope. The proximal descending aorta was then prepared using an external Teflon felt fixed with four internal pledgeted U-stitches. The stent–graft system was then introduced in an antegrade fashion into the descending aorta over the previously positioned stiff guidewire and then released. The side branch was then cannulated and the lower body perfusion was reinitiated. The supraortic vessels were reimplanted, starting from the left subclavian artery. At this point, rewarming was initiated and the proximal repair was finally performed. In case of supra-coronary anastomosis, or patients with previous root replacement, we perform the left carotid and innominate artery reimplantation after the proximal anastomosis at beating heart in order to reduce the myocardial ischemia time.

## Results

The mean age was 59 ± 7.9 years and 5 of them, (62.5%) were male. Aortic aneurysms were the main indication for surgery, with 5 (62.5%) degenerative and 3 (37.5%) post-dissection aneurysms. Hypertension was the most common cardiovascular risk factor (7, 87.5%), and only one patient presented a connective tissue disorder (Loeys-Dietz syndrome). Half of the patients had previously undergone cardiac surgery procedures for previous acute Type A Aortic Dissection.

All patients underwent total aortic arch replacement with the Frozen Elephant Trunk technique, with a zone 2 distal anastomosis and separate reimplantation of the supra-aortic trunks. The brachiocephalic trunk was the most common cannulation site (4, 50%) for cardio-pulmonary bypass, followed by the axillary artery (3, 37.5%) and the ascending aorta (1, 12.5%). Circulatory arrest was performed at 25 degrees Celsius and bilateral antegrade selective cerebral perfusion was always used. Associated procedures were performed in 4 (50%) patients, with Bentall procedure being the most common (2, 25%), followed by coronary artery bypass grafting and aortic valve replacement in one case each respectively.

All 8 patients presented an uneventful post-operative course, with no case of in-hospital mortality nor major complications. In particular, there were no short-term neurological complications, including stroke and spinal cord ischemia. Post-operative outcomes are described in Table 2.

**Table 2:** Post-operative outcomes.

|  | N=8 | N=8 |
| --- | --- | --- |
| In-hospital mortality (n, %) | 0 | 0 |
| Mean ICU stay (days, SD) | 3.5 | 1.4 |
| Mean Hospital stay (days, SD) | 20.1 | 7.4 |
| Neurological complications (n, %) | 0 | 0 |
| Dialysis (n, %) | 0 | 0 |
| GI complications (n, %) | 0 | 0 |
| Respiratory complications (n, %) | 0 | 0 |
| Reopening for bleeding (n, %) | 0 | 0 |
| Laryngeal nerve palsy (n, %) | 0 | 0 |

Mean hospital stay was 20.1 ± 7.4 days.

The main outcome of this observational study is shown in Fig.1, patients induced to moderate hypothermia present low levels of systemic melatonin, being significantly reduced during the t1 condition (P<0.001) and also in the following observational steps (P<0.001 for all comparisons), excluding the G4.

**Figure 1.**
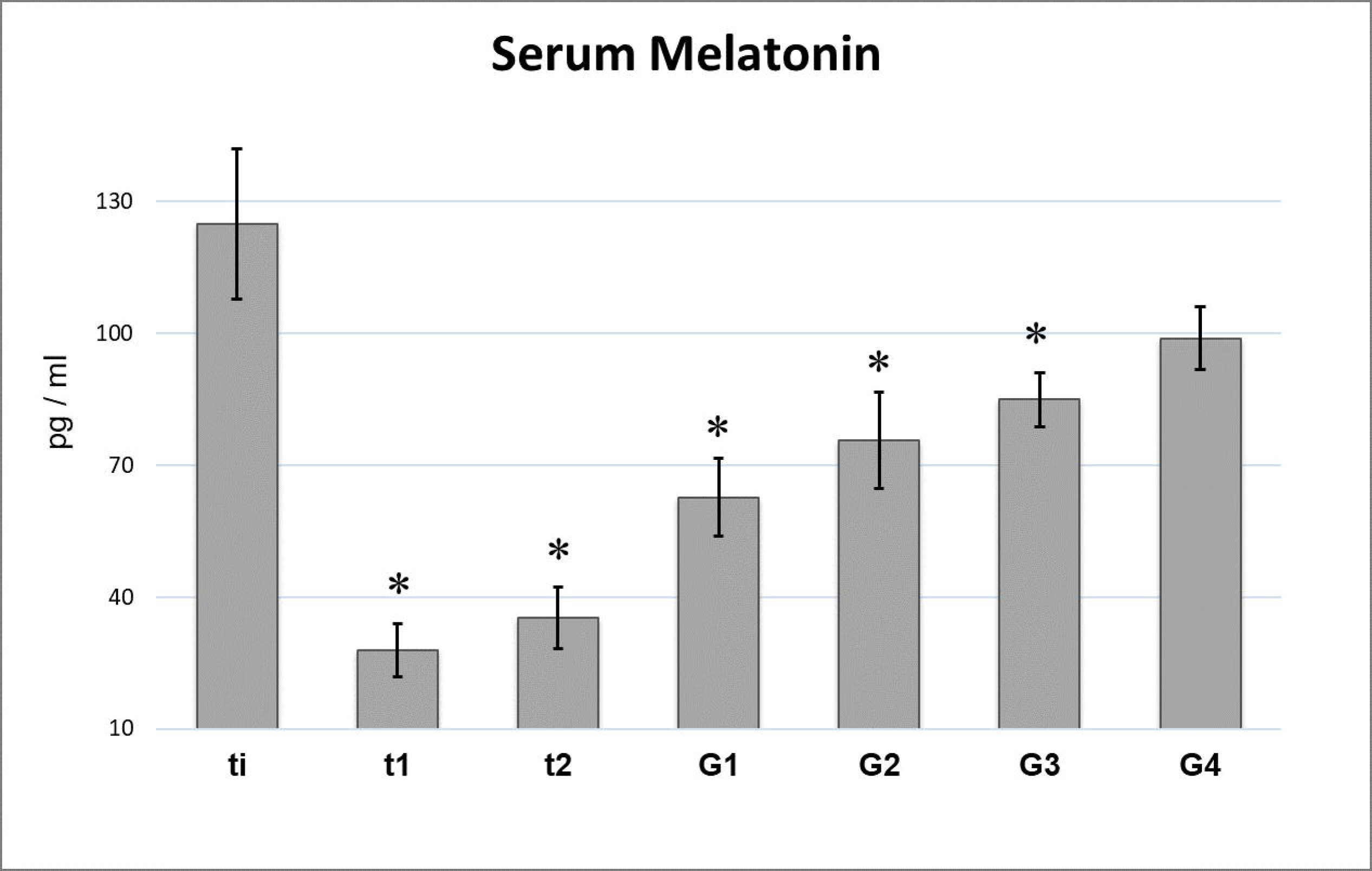
Serum levels of melatonin. Data are expressed as means ± S.E.M., n = 8 (see Methods for details). *: p < 0.05 vs. ti. Observational conditions: ti, induction phase of general anesthesia; t1, target temperature of 25°C; t2, chest closure; G1, first post-operative day; G2, second post-operative day; G3, third post-operative day; G4, fourth post-operative day.

Serum levels of the astrocytic marker S100B (Fig.2) present a huge and significant high peak (P=0.012) in t1, slightly reducing at t3 (P=0.018) and reaching a stable level around double the ti level, however being significantly different in G1 (P=0.012) and G3 (P=0.049). Similarly, serum levels of NSE (Fig.2) present a dramatic and significant rise in t1 condition (P=0.012), but the higher peak value was reached in the t2 sampling (P=0.012). For this marker as well, levels rapidly reduced (see Fig.2) but remaining significantly higher than those in ti in G1 (P=0.012), G2 (P=0.025) and G4 (P=0.036).

**Figure 2.**
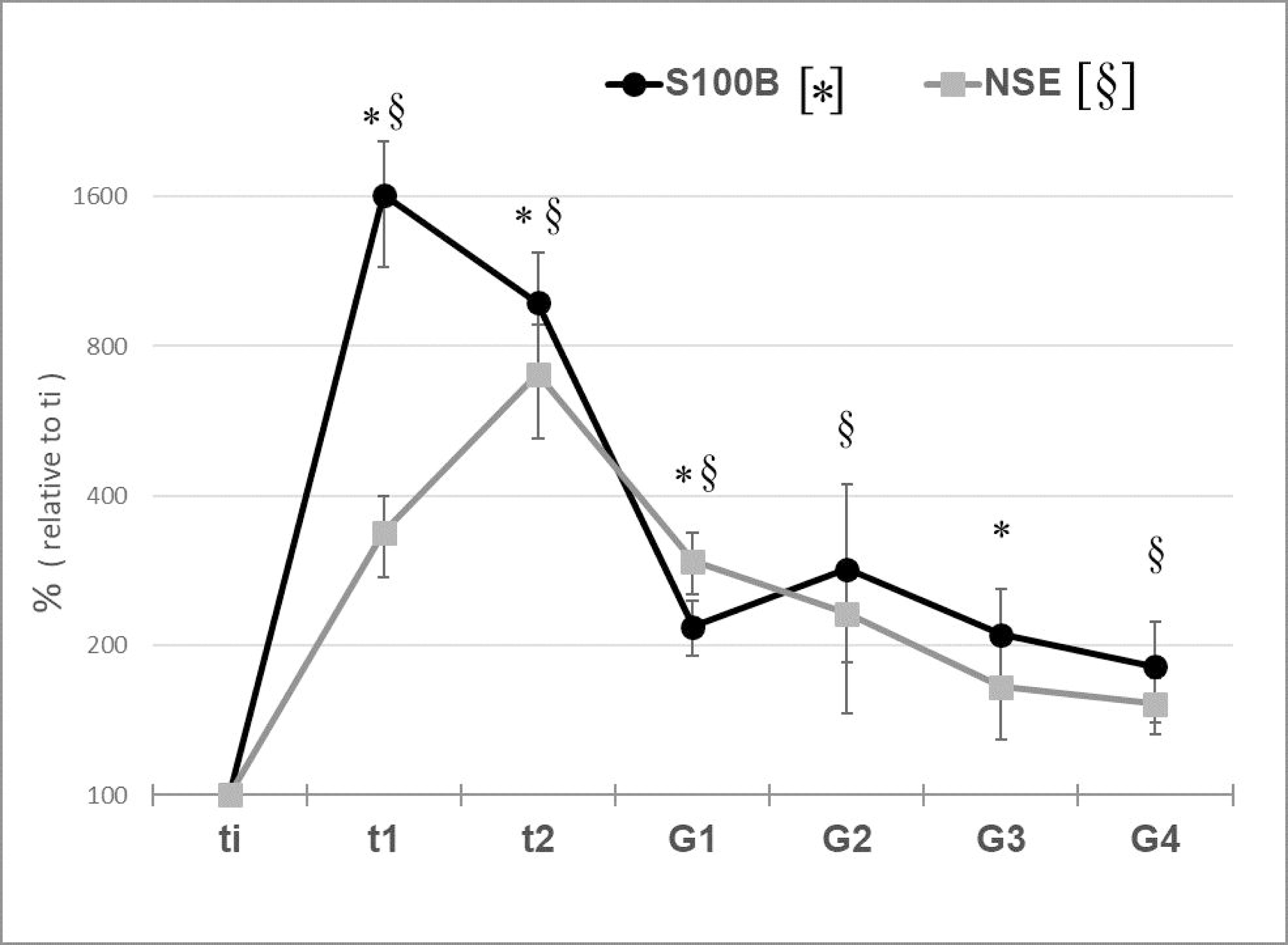
Serum levels of S100B, marker of blood-brain barrier (BBB) integrity, and NSE (neuronal specific enolase), marker of BBB integrity and neuronal damage. Data are shown as percentage in respect to “ti” level and expressed as means ± S.E.M., n = 8 (see Methods for details). *, §: p < 0.05 vs. ti level for S100B and NSE, respectively. Observational conditions: ti, induction phase of general anesthesia; t1, target temperature of 25°C; t2, chest closure; G1, first post-operative day; G2, second post-operative day; G3, third post-operative day; G4, fourth post-operative day.

## Discussions

Results of the present work show that, during the operation, melatonin secretion is strongly inhibited when hypothermia is induced and both S100B and NSE peaked: at the low level of hypothermia and soon at the end of surgery, respectively.

Anesthesia undoubtedly has important effects on the general hormonal pattern (Ivascu et al., 2024), and on melatonin in particular (Guo et al., 2023). However, to the best of our knowledge, nobody measured melatonin secretion during hypothermia, induced in patients while undergoing cardiac surgery. It is well-established that the disruption of melatonin secretion represents one of the main parameters leading to common circadian disturbances in post-operative recovery (Guo et al., 2023), and this may explain the lower levels we found during the first 72h of recovery from surgery (see Fig.1). However, the most peculiar result in serum melatonin we obtained is that relative to the hypothermic condition. As reported by Reber and colleagues (1998), during anesthesia melatonin plasma level is slightly affected and this is in contrast to what was found here: serum melatonin was dramatically low during the hypothermic period of the surgical procedure.

Neurological complications in cardiac surgery, and aortic arch procedures with hypothermic circulatory arrest in particular, have long been evaluated. These include not only stroke and paraplegia, but also more subtle post-operative cognitive disorders such as delirium for which the pathophysiological basis is yet to be clear. Post-operative delirium is strongly linked to alteration of the circadian rhythm disruption, as is melatonin secretion (Kim et al., 2023; Fan et al., 2024; Itting et al., 2025).

The effect that hypothermia may have on melatonin levels was precisely the focus of the present working hypothesis, built on the previous finding that, in rodents, a reversible hypothermic condition pharmacologically induced, the ST, was able to trigger a molecular neuroprotective mechanism, probably linked to the high levels of melatonin found (Squarcio et al., 2023). Then, present data disprove that hypothesis.

Hence, neither surgery nor hypothermia alone apparently explain the important drop of serum melatonin we found. It follows that the present results may be interpreted as due to the concomitant occurrence of the two factors.

However, the drop of melatonin we observed in the present work, together with our preclinical data (Squarcio et al., 2023), also suggest a possible way to stimulate neuroprotection and probably improve the clinical outcome of these patients: by administering melatonin to patients, at the beginning of the hypothermia induction. As a matter of fact, melatonin is a well-known safe molecule, approved by the US Food and Drug Administration (FDA) as a “dietary supplement”, normally used to cope with mild sleep disturbances or jet-lag circadian disruption. In addition, melatonin has numerous well described effects as antioxidant and anti-inflammatory agent (Kücükakin et al., 2009), also showing promising results when given in peri-operatory conditions and in particular in relation to neuropsychiatric outcomes in cardio-pulmonary surgery (Artemiou et al., 2015; Shin et al., 2024). Therefore, since at physiological temperature melatonin appears to have good peri-operatory protective effects, mainly acting on the regularization of the circadian cycle, our pre-clinical indications support the possibility that, if the treatment is given at around 25°C of brain temperature, this may have even stronger neuroprotective effects.

The role of S100B and NSE as serum biomarkers of the BBB integrity and neuronal damage, respectively, is very well established (Marchi et al., 2004; Hajduková et al., 2015; Friis et al., 2022; Sandroni et al., 2023). The rise of serum level of the astrocytic protein S100B is linked to a BBB higher leakage, while NSE, being a metabolic enzyme rich in neurons, when found in serum is a sign of possible neuronal damage (Marchi et al., 2004). It is worth noting that the levels of these biomarkers, apparently, are not modified by hypothermia (Pfeifer et al., 2014). Present results show that S100B peaks during hypothermia, that is also concomitant with the ongoing cardiopulmonary bypass, and persisted until the end of the surgery, then reducing but maintaining statistically significant high levels until G3. This was expected, since a disruption of BBB following cardiopulmonary bypass has been already described (Abrahamov et al., 2017). As specified by different works in literature, in extreme cases, like following serious neural insult due to cardiac arrest, serum NSE peaks at 24-72 h after the insult, and significant high levels of this biomarker can easily be found at 4-5 days and beyond (Pfeifer et al., 2014; Gul et al., 2017; Sandroni et al., 2023). Then, our results, shown in Fig.2, apparently indicate that the present surgery procedure induces some acute neural suffering, but rapidly solving in a very few days in respect to what found in more significant neural injuries (Pfeifer et al., 2014).

Considering these data together and the very high level of melatonin safety, this study may open new avenues to treat subjects in which the hypothermia is normally used for aortic arch surgery. Administering melatonin while the patient is hypothermic should compensate the serum melatonin lack we observed. Melatonin, by acting at low temperature and easily passing the BBB due to its temporary inefficiency, should trigger the molecular neuroprotective mechanism we observed in the pre-clinical study by Squarcio and colleagues (2023), acting as a long-desired safe GSK3β inhibitor to contrast neuronal suffering (Beurel et al., 2015). Consequently, this easy treatment may improve the neurological outcomes of this kind of surgery (i.e., lowering serum NSE levels), with virtually no aversive effects.

## Data Availability

All data produced in the present study are available upon reasonable request to the authors

## Abbreviations

BBB: blood-brain barrier
G1: first post-operative day
G2: second post-operative day
G3: third post-operative day
G4: fourth post-operative day
GSK3β: glycogen-synthase kinase
3β NIRS: near infrared ray spectroscopy
NSE: neuronal specific enolase 2
S100B: astrocytic protein, peripheral marker of BBB integrity
ST: synthetic torpor
t1: target temperature of 25°C
t2: chest closure;
ti: induction phase of general anesthesia

## Research data for this article

Research data are available upon reasonable request to the corresponding author.

## Acknowledgments

We would like to thank the company Eurosets S.r.l. allowing us to use serum samples taken for a study they conducted, without asking any economical or scientific restrictions nor interfering with our results or implication discussed in the present work.

## CRediT authorship contribution statement

**Leone Alessandro:** Conceptualization, Data curation, Investigation, Formal analysis, Project administration,Writing-original draft, Writing-review & editing; **Occhinegro Alessandra:** Investigation, Formal analysis; **Piscitiello Emiliana:** Investigation, Formal analysis; **Paterini Paola:** Investigation; **Bugani Simone:** Supervision; **Mariani Carlo:** Investigation; **Nocera Chiara:** Investigation; **Hitrec Timna:** Investigation, Writing-review & editing; **Amici Roberto:** Supervision, Writing-review & editing; **Pacini Davide** Conceptualization, Project administration, Supervision, Writing-review & editing; **Luppi Marco:** Conceptualization, Data curation, Formal analysis, Funding acquisition, Project administration, Supervision, Visualization, Writing-original draft, Writing-review & editing.

## Ethics approval statement

The study was observational and conducted according to the guidelines of the Declaration of Helsinki and approved by the Institutional Ethics Committee: *Comitato Etico - Area Vasta Emilia Centro* (CE-AVEC). Protocol approval code 127/2022/Oss/AOUBo.

## Patient consent statement

Written informed consent was obtained from all patients.

